# Artificial intelligence assisted standard white light endoscopy accurately characters early colorectal cancer: a multicenter diagnostic study

**DOI:** 10.1101/2020.02.21.20025650

**Authors:** Sijun Meng, Yueping Zheng, Ruizhang Su, Wangyue Wang, Yu Zhang, Hang Xiao, Zhaofang Han, Wen Zhang, Wenjuan Qin, Chen Yang, Lichong Yan, Haineng Xu, Yemei Bu, Yuhuan Zhong, Yi Zhang, Yulong He, Hesong Qiu, Wen Xu, Hong Chen, Siqi Wu, Zhenghua Jiang, Yongxiu Zhang, Chao Dong, Yongchao Hu, Lizhong Xie, Xugong Li, Jianping Jiang, Huafen Zhu, Wenxia Li, Zhang Wen, Xiaofang Zheng, Yuanlong Sun, Xiaolu Zhou, Limin Ding, Changhua Zhang, Wensheng Pan, Shuisheng Wu, Yiqun Hu

**Author notes:** **Correspondence to:** Changhua Zhang: Center for Digestive Disease, the Seventh Affiliated Hospital of Sun Yat-sen University, No. 628, Zhenyuan Rd, Guangming (New) Dist, Shenzhen, Guangdong Province, P. R. China, 518107., Shuisheng Wu: College of Pharmacy, Fujian University of Traditional Chinese Medicine, No.1 Qiuyang Road, Shangjie University Town, Fuzhou, Fujian, P.R. China, 350122., Wensheng Pan: Zhejiang provincial people’s hospital, People’s Hospital of Hangzhou Medical College, No. 158 of Shangtang Road, XiaCheng District, Hangzhou, Zhejiang Province, P. R. China, 310014., Yiqun Hu: Department of Gastroenterology, Zhongshan Hospital Xiamen University, 201 Hubin South Road, Xiamen, Fujian Province, P. R. China, 361004.

## Abstract

Colorectal cancer (CRC) is the third in incidence and mortality^1^ of cancer. Screening with colonoscopy has been shown to reduce mortality by 40-60%^2^. Challenge for screening indistinguishable precancerous and noninvasive lesion using conventional colonoscopy was still existing^3^. We propose to establish a propagable artificial intelligence assisted high malignant potential early CRC characterization system (ECRC-CAD). 4,390 endoscopic images of early CRC were used to establish the model. The diagnostic accuracy of high malignant potential early CRC was 0.963 (95% CI, 0.941-0.978) in the internal validation set and 0.835 (95% CI, 0.805-0.862) in external datasets. It achieved better performance than the expert endoscopists. Spreading of ECRC-CAD to regions with different medical levels can assist in CRC screening and prevention.

## Introduction

Colorectal cancer (CRC) is one of the most common malignant tumors in the globe^4^. “Cancer Statistics, 2020” indicates that CRC ranks the third in incidence and mortality^1^. Accurate identification and treatment of early CRC have significantly reduced incidence and mortality^5^. However, early CRC is generally asymptomatic, and coloscopy allows direct visual inspection of the intestinal tract and same-session detection, biopsy, and subsequent removal of disease lesion. Several countries recommend that adults over the age of 50 should regularly screen CRC by coloscopy^6,7^. While coloscopy is a kind of expert dependent examination, which can lead to missed diagnosis and misdiagnosis due to various causes, such as the experience of endoscopist, the interference during operation and inevitable interval of eye movement^8^. Computer assisted diagnose (CAD) system can solve above-mentioned problems.

In recent years, some Artificial Intelligence (AI)-based CAD systems have come into the pre-clinical application stage. However, the limitations of the research design (i.e. lack of external validation, not solving the critical clinical problems) lead to the consequence that the existing assistances are not really suitable for clinical practice directly^9,10^. In the field of gastrointestinal tumor, most studies focus on the detection of polyps and adenomas^11,12^, the staging of advanced cancer^13^ and the optimizing of endoscopic operation^14,15^. Effects of these CAD systems in decreasing mortality rate of CRC need to be proven^16^ by more clinical trials. Early gastrointestinal cancer diagnosis system that can directly decrease the occurrence of CRC is limited^17^, because the quantity of images from a single medical center is insufficient to support the establishment of the algorithm. Therefore, we aim to establish an early CRC CAD and malignant potential characteristic system (herein referred to as ECRC-CAD) based on standard white-light endoscopy (WLE), which can be clinically applied to different regions. In this multi-center diagnostic clinical research, we establish an AI assisted early CRC diagnosis model using deep learning, which is based on images of different quality endoscopy from hospitals in different medical levels (i.e., rural, municipal, provincial) in China. Furthermore, ECRC-CAD can identify intraepithelial neoplasm (IN), which is one of the most typical malignant precancerous conditions that can be differentiated by using image-enhanced endoscopy (IEE) and diagnosed by histology test. The clinical applicability of this model has been verified in the internal and external datasets collected retrospectively and anther internal dataset collected prospectively.

## Methods

### Study design and participants

This study was a multi-center clinical diagnose research, which was completed in five hospitals at different medical levels across three provinces of China.

The workflow is shown in Figure 1. The first part is a case-control, retrospective research. All patients who underwent coloscopy examination excepting for repeated examination in four hospitals from January 2016 to July 2019 are enrolled, including Zhongshan Hospital of Xiamen University (XMZSH) in Fujian, Zhejiang Provincial People’s Hospital (ZJPH) in Zhejiang, the Seventh Affiliated Hospital of Sun Yat-sen University (SYSUSH) in Guangdong, and Pucheng County Traditional Chinese Medicine Hospital (PCTCRMH) in Fujian. Among them, SYSUSH started operation in July 2018. Patients who were diagnosed with early CRC by histopathological examination were selected. Pathological diagnosis was referred to the 4th WHO Classification of Tumors of Digestive System^18^. Early CRC in this article referred to precancerous conditions, in accordance with the requirements of the international standard. Patients with progressive colorectal cancer were excluded. Images of early CRC confirmed to the database’s including criteria (see in below) were included into case-control study, and were randomly assigned to the training set, evaluation & tuning set, and internal testing set in the ratio of 8:1:1. All images of intraepithelial neoplasm (referred as IN-ECRC) were included to case group, and other early CRC without intraepithelial neoplasm (referred as NIN-ECRC) were randomly sample in corresponding quantity included to control group. ECRC-CAD was constructed and optimized using the first two data sets. To verify the veracity and clinical applicability of the model, we used validation datasets including images from the above internal testing set and another external testing set, which are consisted of images are according to the above criteria from Tongxiang Hospital (TXH) in Zhejiang. The second part is a prospective research. The identification model officially completed the detection and model optimization of the above samples in September 2019, and was connected to the above five hospitals through the Internet. Then we prospectively collected the AI recognition results of all endoscopic center images in October 2019 and the first half of November 2019 from the above five hospitals, and extracted some endoscopic images of IN-ECRC and NIN-ECRC for senior doctors to identify.

**Figure 1.**
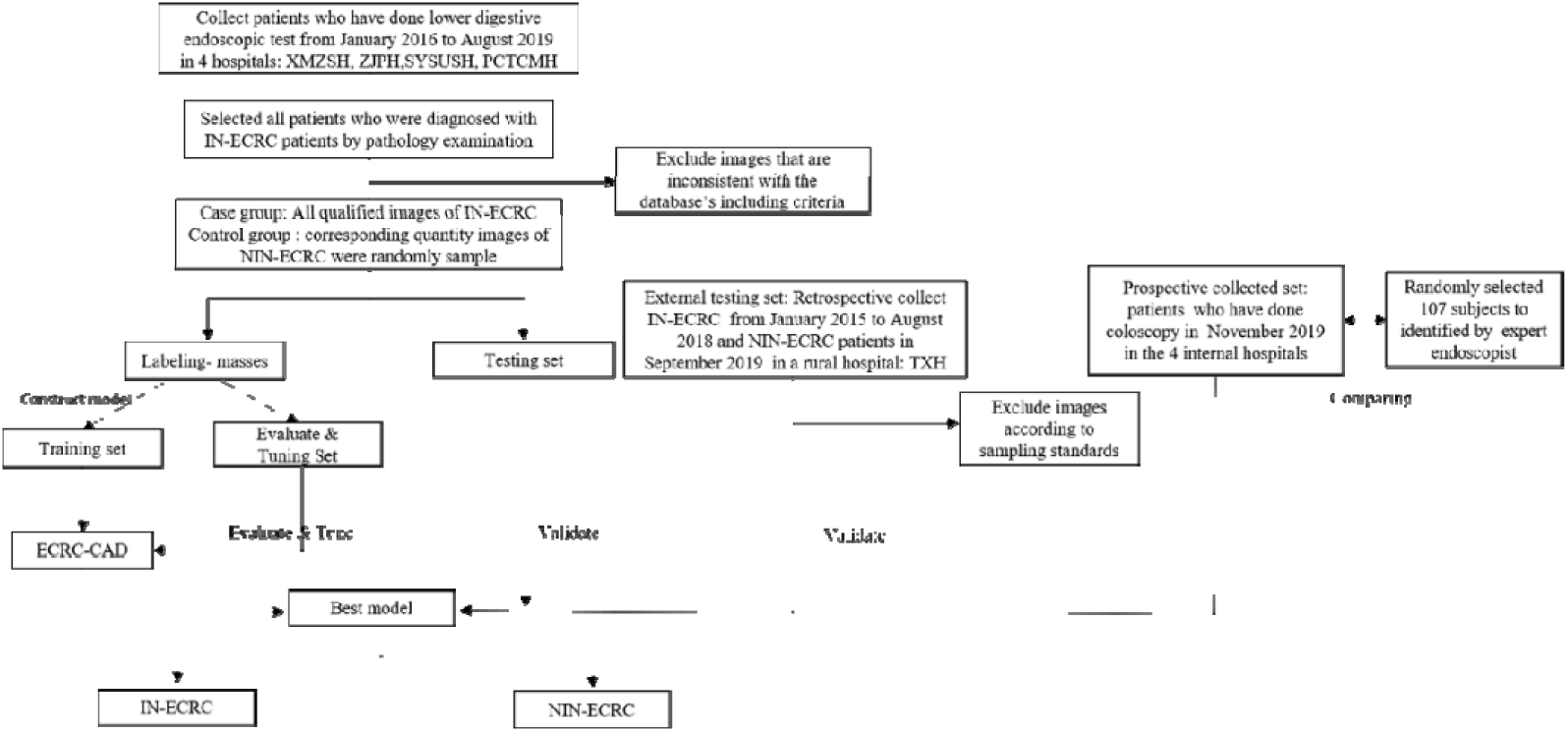
Workflow of establishment and valuation of ECRC-CAD. The flow chart shows the construction and validation process and committed steps of ECRC-CAD.

Each participating institute has got ethical approval by independent ethics Committee according to the Helsinki declaration. The informed consent of retrospective cohort was exempted by the institutional review boards, causes it’s the secondary use of clinical data without any intervention. In the prospective study, each patient was informed and signed up informed consent.

### Endoscopy and early CRC’s image database

Endoscopic images of five hospitals were captured by EC590WM, EC-600WM, EC-L590Z (WFujinon Corp, Saitama, Japan); V-70, PCF-H260AZI, CF-H260AI, CF-H290I, CF-HQ290I (Olympus Medical Systems, Tokyo, Japan); EC34-i10M,EG2990Z I (PENTAX medical, Tokyo, Japan). Each image of patients with early CRC was sequenced according to anatomical location, and screened by three endoscopy physicians. Exclusion criteria include: 1. The endoscopic anatomical location was not consistent with the pathological report, 2. Pictures were taken under IEE, 3. Pictures without any early CRC lesion, 4. Pictures was not confirmed to the sampling standard^19^ summarized by our research team. All images in the database present at least one lesion, and multiple images might be taken for the same lesion, including differences of angle, distance, and extension of the intestinal wall. Images were saved in a jpeg format.

### Image Labeling

Two senior endoscopists with more than 10 years of clinical experience, four residents and two assistants carried out image selection and labeling with blind method. The doctors were divided into three groups randomly, and at least one senior doctor in each group. The deep learning marking tool LabelMe20 of Anaconda was used to mark the outline of disease lesion manually. Two physicians in the same group worked together. One was mainly responsible for drawing, and the other was mainly responsible for checking. Only when two physicians from the same group reached a consensus, the labeling of the image could be finalized. After the first identification by ECRC-CAD, the misdiagnosed images were double-checked so as to ensure the correctness and rationality of the results.

### Construction and optimize of algorithm

The ECRC-CAD’s algorithm was based on the concept of full convolution networks^21^ (FCN), which is composed of classification module and post-processing module (Supplements p1-p2). The pattern diagram was shown in Figure 2. Comparing with the classical CNN, this method can be adapted to inputs with any size. In order to get an accurate boundary, we precisely outlined disease lesion of images in establishing sets. The model contains one input and one output. The input of the model was the image that contains the lesion. The output implemented a segmentation task that each pixel is classified as background, intraepithelial neoplasm or other early CRC. Intraepithelial neoplasm was abbreviated as “IN-ECRC”, and the other early colorectal cancer neoplasm was abbreviated as “NIN-ECRC”. The labeling from histopathological examination was adopted as the gold standard in training ECRC-CAD.

**Figure 2.**
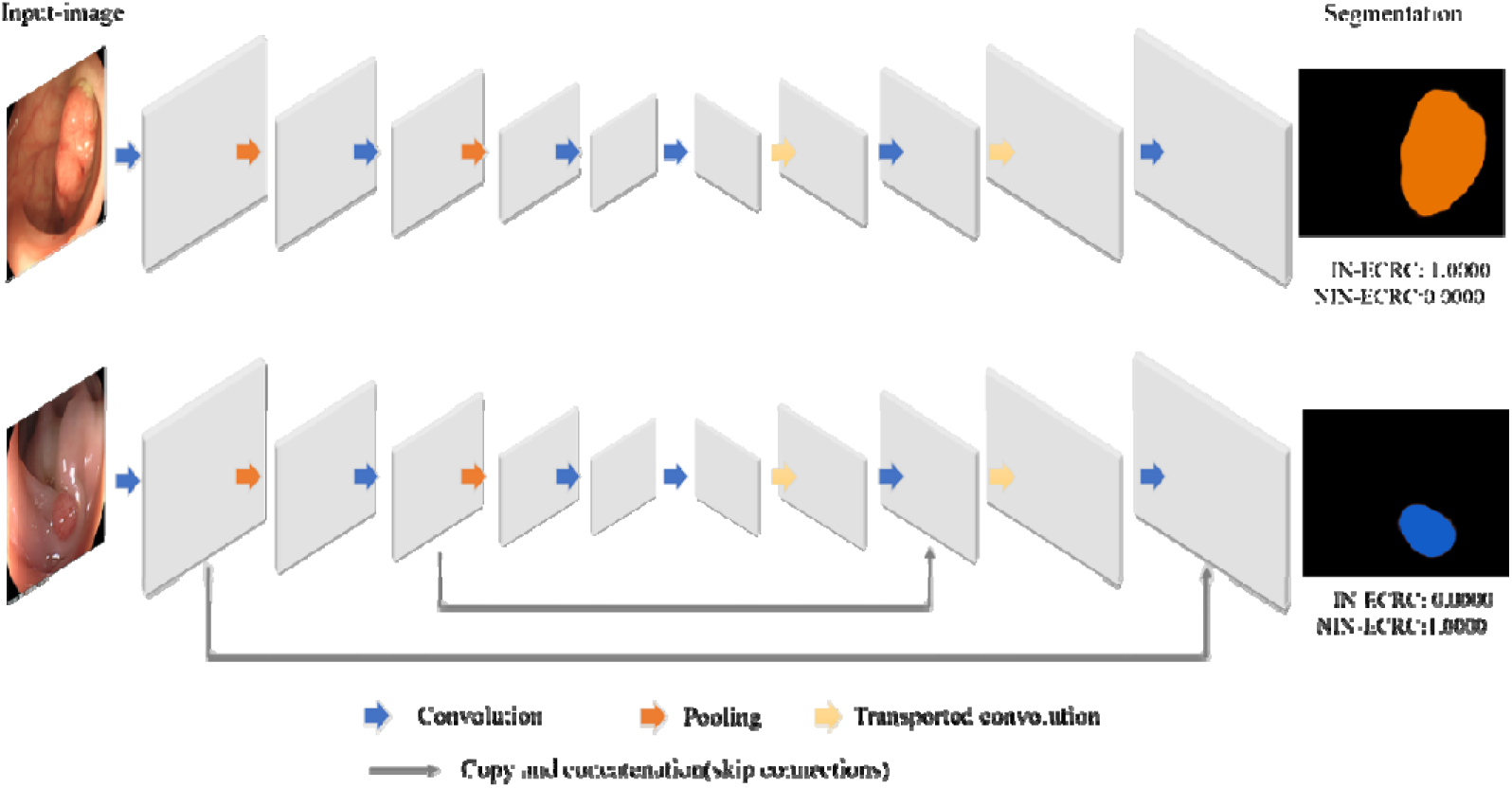
Diagram of ECRC-CAD. Algorithm of ECRC-CAD is based on FCN8. Score of IN-ECRC+ score of NIN-ECRC=1, If score of IN-ECRC >score of NIN-ECRC, we recognize it as IN-ECRC, if score of NIN-ECRC>score of IN-ECRC, we recognize it as NIN-ECRC.

**Figure 3.**
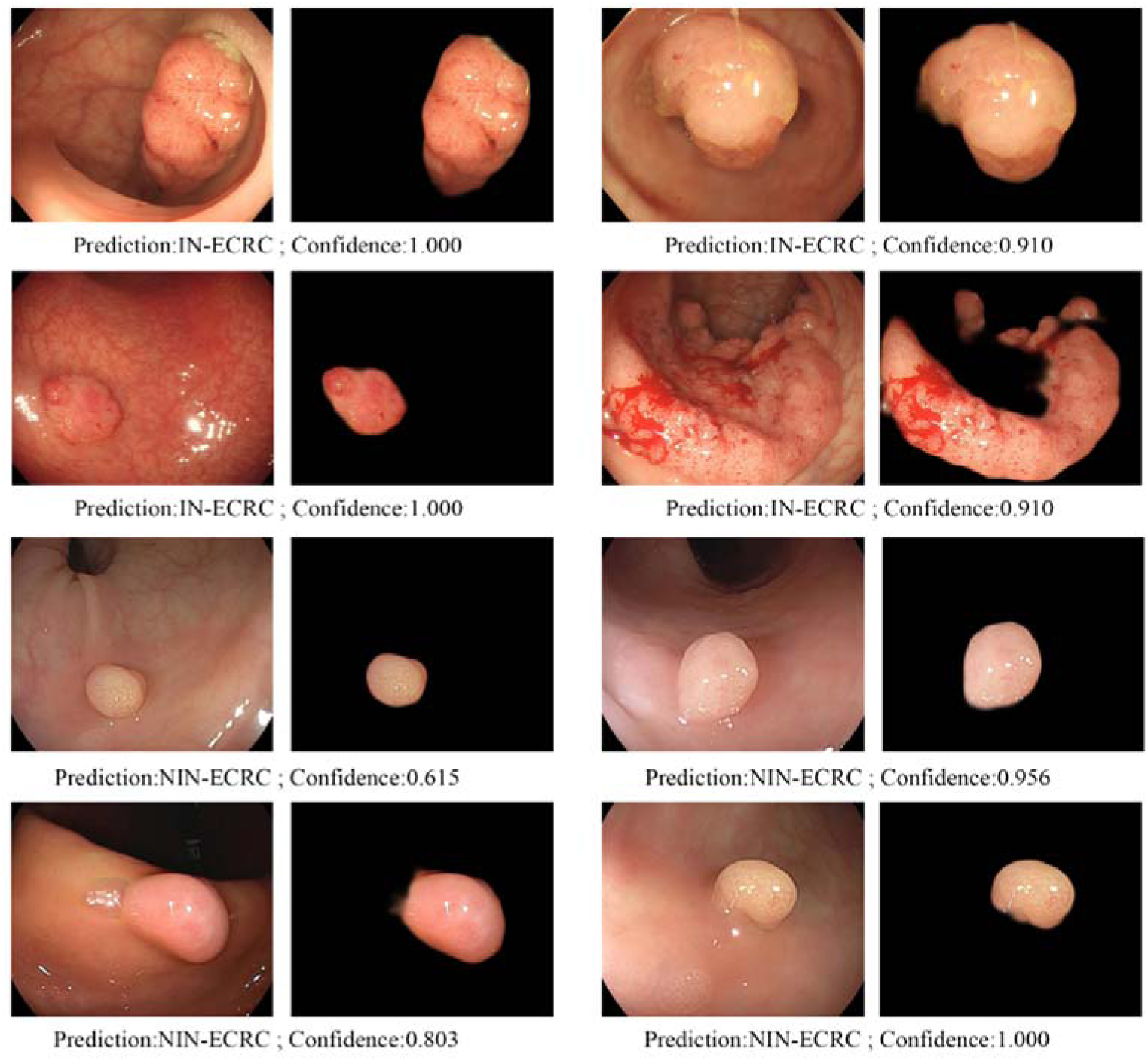
Demonstration of ECRC-CAD’s extraction and characterization results. The left image of each pair is the original image under the endoscope, the right image of each pair is the disease lesion extracted by ECRC-CAD. Predict result was listed under each pair of images. The “Prediction” result represents the type of extracted lesion identified by ECRC-CAD, the “confidence” represents the score that were calculated by ECRC-CAD.

The implementation of AI-assisted precursor lesion identify model was fulfilled by using Python 3.7 (Python Software Foundation; www.python.org/).

### Algorithm testing and outcome measurement

Three validation datasets were used to verify ECRC-CAD, which include internal datasets, external datasets, and prospectively collected datasets. In the internal and external validate datasets, the performance of ECRC-CAD was compared with the pathological golden standard of the corresponding tissues, and calculated statistic indicators on the individual images. When FCN detects early CRC from the input data of the validation image, the model outputs the lesion name. When the output result of the model is consistent with the pathological result, it is considered to be correct. In the prospective dataset, the performance was compared with histopathological result and expert at the same time. We randomly selected a subset of patients’ images with histologically confirmed early CRC from the prospective collected datasets. Two experienced endoscopists were asked to complete independently the same test images together. These images’ histology diagnosis is blinded to the two endoscopists. The diagnosis results of ECRC-CAD and experts were taken the patient as the unit, which is according with clinical requirements.

Sometimes, it’s difficult to identify the whole images of disease lesion caused by the image quality and angle. When the model can identify even one picture of the cancer, it is considered as correct.

### Statistical analysis

In this study, descriptive analysis was used to describe the baseline characteristics of the enrolled subjects. Continuous variables were expressed by median (interquartile interval), dichotomous variables and grade variables that expressed by percentage. In order to evaluate the authenticity of ECRC-CAD, the diagnostic evaluation method was adopted. The evaluating indexes included Sensitivity, Specificity, Condolence, Positive predictive value (PPV), and Negative predictive value (NPV). According to the above indexes, receiver operating characteristic (ROC) of different subgroups were drawn, and the area under the ROC curve (AUC) was calculated to evaluate the classification effect of this model on different subjects. More details are in the supplements p3. All statistical analyses in this study were based on the two-sided test 0.05 and performed through R software version 3.6.0 (R Development Core Team).

## Results

### Patient and image dataset information

More than 6.2 million patients have done lower gastrointestinal tract in XMZSH, ZJRMH, SYSUSH, and PCRMH from January 2016 to August 2019. Almost 40% percent without any disease lesion were excluded. All participants diagnosed with IN-ECRC were enrolled in the model establishing sets, after quality control, 1923 images were enrolled, participants with NIN-ECRC corresponding in number of IN-ECRC were randomly extracted from other patients. The ratio of male to female is about 2.08:1. Age ranges from 20 to 86, and represent the right-skewed distribution (median age is 53.5). The baseline information of the establishing cohort and external cohort were listed in Table 1. Finally, in the stage of model establishment, 1923 images included in the case group, and 2467 images were included in the control group. In the model optimization stage, 490 images were included. In the verification stage, 189 images and 247 images were respectively collected in the above four hospitals as internal datasets. External datasets, including 960 images from 134 patients, were used to contract and test ECRC-CAD. The prospectively collected datasets enrolled 672 patients, including 319 IN-ECRC and 353 NIN-ECRC patients.

**Table 1.** Baseline information.

|  |  | Establish cohort |  |  | Validate cohort |  | Expert<br>endoscopists<br>(unit: person) |
| --- | --- | --- | --- | --- | --- | --- | --- |
|  |  | Establishing set<br>(unit: image) | Evaluate & Tuning<br>set (unit: image) | Internal testing set<br>(unit: image) | External testing set<br>(unit: image) | Prospective testing set<br>(unit: person) |  |
| Sum of subjects |  | 3464 | 490 | 436 | 674 | 454 | 107 |
| Stage | IN-ECRC | 1491 | 243 | 189 | 674 | 98 | 63 |
|  | NIN-ECRC | 1973 | 247 | 247 | NaN | 353 | 44 |

### Performance of the precancerous detection model

ECRC-CAD output results within 0.16 seconds and marks the boundary of disease lesion. The diagnosis result of each image was listed in supplementary material (eTable 1). The diagnostic accuracies of the internal validation dataset were 0.963 (95% CI, 0.941 to 0.978). External testing set only includes only IN-ECRC, which causes pathologists found that diagnostic criteria of NIN-ECRC in TXH are not based on the 4th WHO Classification of Tumors of Digestive System. The sensitivity of ECRC-CAD in THX is 0.835 (95% IC, 0.805 to 0.862). The four indicates of the internal validation dataset is higher than 0.93. Similarly, a high AUC value (0.939) was also observed in the internal datasets (Figure 4). In the prospective study, the index of diagnosis accuracies was calculated in human units, which is more suitable for clinical needs. High accuracy (0.885, 95% CI, 0.859 to 0.907) was observed. The sensitivity and PPV were more than 0.90, and specificity and NPV were more than 0.85. The causes of misidentification are similarities, including incomplete exposure of disease lesions, concomitant other diseases, and some normal structures. The PPV (0.609, 95% IC, 0.454 to 0.745) and NPV (0.426, 95% IC, 0.303 to 0.559) were significantly lower for the expert endoscopists when compared with the AI-assisted model (*P*<0.0001). Sensitivity and specificity between the endoscopic experts and the ECRC-CAD were visible.

**Figure 4.**
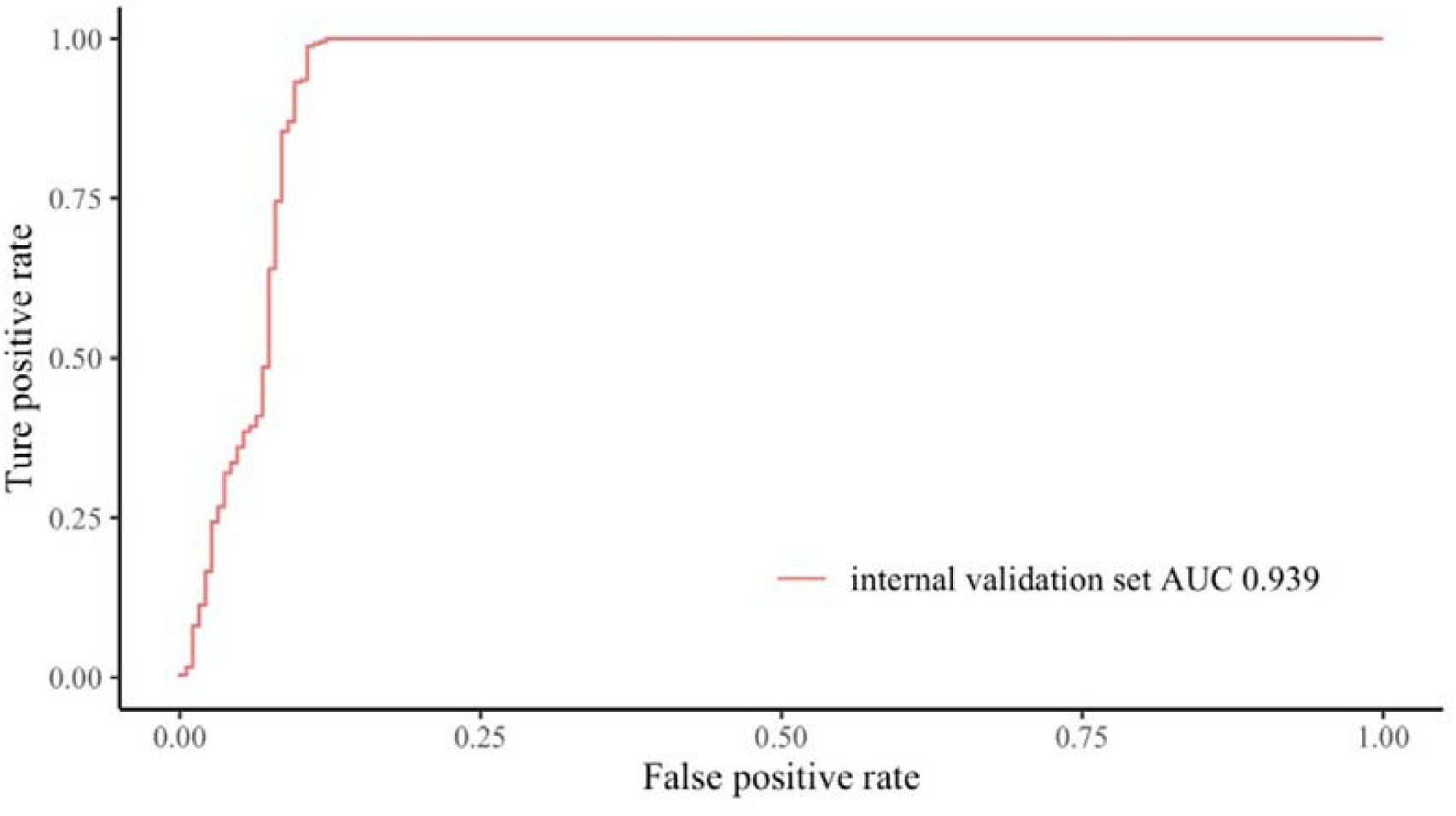
Receiver operating characteristic curves of ECRC-CAD. ECRC-CAD performance on the internal validate sets (436 images). AUC indicates area under the receiver operating characteristic curve.

This model has been successfully embedded in the above five hospitals, which is connected to the endoscopically connected computers through the Internet. Doctors and clinicians can identify the routine screenshots during the operation. The results of model recognition are displayed in the upper right corner of the screen in real time. In order to benefit more people, ECE-CAD is now available at: http://123.57.221.125/#/home. Home page is on display in Supplementary material (eFigure 2). Endoscopists can upload endoscopic images of suspected early CRC, and the system will give judgment in real time. Furthermore, we provided a set of WLE images of early CRC through open ports, which can help inexperienced endoscopist to familiar with it. The openness of this recognition model can also help endoscopists in less developed areas or those with low seniority to learn the recognition of early CRC (eFigure 3).

Both Asian and Weatern researchers consider that specific gross morphology, mucosal surface structure and microvascular structure or multi-changes of neoplasm have corresponding molecular characteristics^22^. We analyzed the microstructure feature of WLE, IEE in the same lesion taken from the same angle, and the association with features of H&E stained pathological image (eFigure 4). Combined with the published results (eTable 2), we proposed that the application of endoscopic images to predict molecular characteristics through endoscopic or pathology images in real-time is the future direction of “Image genomics”^23^.

**Table 2.** Performance of ECRC-CAD in different validation dataset and expert endoscopist in distinguishing IN-ECRC and NIN-ECRC

|  | Internal testing set | External testing set | Prospective testing set | Expert endoscopist | ECRC-CAD vs<br>Endoscopist* |
| --- | --- | --- | --- | --- | --- |
| Accuracy (95% CI) | 0.963 (0.941-0.978) | 0.835(0.805-0.862) | 0.885 (0.859-0.907) | 0.505 (0.411-0.598) | *P*<0.0001 |
| Sensitivity (95% CI) | 0.937 (0.889-0.965) | 0.835(0.805-0.862) | 0.915 (0.878-0.942) | 0.444 (0.321-0.575) | *P*<0.0001 |
| Specificity (95% CI) | 0.984 (0.956-0.995) | NaN | 0.858 (0.817-0.892) | 0.591 (0.433-0.733) | *P*<0.0001 |
| PPV (95% CI) | 0.978 (0.941-0.993) | NaN | 0.854 (0.811-0.887) | 0.609 (0.454-0.745) | *P*<0.0001 |
| NPV (95% CI) | 0.953 (0.917-0.974) | NaN | 0.918 (0.882-0.944) | 0.426 (0.303-0.559) | *P*<0.0001 |

\*McNemar's test

## Discussion

Overall, 1680 images of IN-ECRC and 2220 images of NIN-ECRC were used to establish and optimize ECRC-CAD. In order to ensure the clinical application of the model in underdeveloped regions, these images were collected from different medical level hospitals in China. Review published articles, it’s the largest, multi-source endoscopic image database used to establish AI aided diagnose system in globe^24^. Furthermore, three clinical validation studies were conducted on the immediate clinical application value of ECRC-CAD, which showed high accuracy in the internal dataset (0.963). Liu et al^25^ systematically reviewed the in-depth learning performance of diagnosis of disease from medical imaging. Of the 82 eligible researches from 2012 to 2019, only 25 have external validation. We avoided the issues mentioned above in this meta-analysis. The displayed ECRC-CAD can be extended to the external dataset from another rural hospital with a sensitivity of 0.835. This verification cohort could avoid highly accurate prediction which are caused by excessively spurious associations with the known outcome. However, case-control would not be representative of the general screening population^26^, and the predictor may not be directly applied to clinical practice. In this study, we validated ECRC-CAD in a prospective cohort, and ECRC-CAD and experts identified the disease’s nature in real-time, respectively. The result showed that ECRC-CAD maintained perfect accuracy and specificity in prospective study, even better than expert endoscopist.

In clinical daily work, ECRC-CAD can assist endoscopist through increasing early CRC’s detection rate, decreasing missed diagnosis rate (Figure S5). Endoscopy and clinical practice of operator have a great influence on the detection and treatment of early cancer. A prospective study of UK^27^ shows that the good performance of operating require skilled intubation operation and independent practice. While endoscopist in developed region can get more information about microstructure under IEE, which can increase diagnosis rate of early CRC by 10%-20%^28^ comparing with WLE. However, early screening of neoplasm is mostly carried out in community hospitals with relatively low medical level, and the endoscopic equipment is relatively backward. Early CRC detection rate of ECRC-CAD has the same performance with IEE. Using ECRC-CAD enable people in lower medical level regions to get similar screening efficiency compared with subjects in developed regions.

Apart from assisting clinical decision, AI can make clinical path optimization possible^23,29^. The introduction of colorectal endoscopic submucosal dissection (ESD) increased the complete resection rate of early CRC^30^ with minimally invasive trauma and low local recurrence possibility. High-grade IN without invasion or only superficial submucosal infiltration diagnosed by pathology examination is an indicator for ESD. The interval time between first endoscopic biopsy and pathology diagnosis is usually one week. At present, multi classification based on IEE^31–33^ has been widely used to predict the risk of malignancy and invasiveness of CRC. Neoplasm confirm to indicators of malignant aggressiveness subtype according to these classifications can be removed during first endoscopic operation. It saved medical resources, and more important, absence of IEE or experts may increase metastasis of implanted cancer cell in natural cavity^34^ due to incomplete resection. But there’re some limitations: Kudo, Sano classification is based on single observation index of pit or vessel pattern, NICE classification combining with gland and capillary pattern cannot differentiate IN from NIN, and all classifications need expert practice and IEE. ECRC-CAD can identify IN under WLE within 0.16 second, which make up for the above deficiencies of present classification.

WL Bi^23^ mentioned that characterization and monitoring are the future direction of AI in medical image. ECRC-CAD have realized characterization of precancerous lesion, and it can steadily monitor the progression of CRC without interference. However, with accumulation of medicine data preternaturally over time, AI will add rich layers of intelligence through combining molecular and pathological information with image-based findings. “Image genomics”^23^ is the key to realize it, referring to the association of imaging characteristics with biological data, including specific molecular markers that are mainly based on immunohistochemical, or sequencing technology that are developed to direct personalized treatment planning. In lung cancers, researchers have established algorithm to predicted EGFR genotype using computed tomography image (MRI). Can endoscopic images be used in image genomics? We reviewed all researches of surface microstructure under endoscopy and molecular characteristics associated with colorectal neoplasms. These results indicate that that the AI have the potential of identify phenotypic and molecular specific pathological changes under WLE, causes of AI can extract a large number of features that cannot be observed by the human naked eyes. The development of predicting multigroup tumor molecular markers through endoscopic images will paved the way for the future “one-step” clinical workflow with AI interventions.

The insufficiencies in this study includes: 1. ECRC-CAD can’t increase adenoma detection rate (ADR), because it’s based on static image taken by endoscopist. We’ve explored a CAD based on video, and the clinical validation is processing. 2. The external validation set is not included with NIN-ECRC. We are reassessing this part of cases by two expert histopathologists, to add a complete evaluation to this model. 3. Sample of high-grade IN is limited for making differentiation of ECRC-CAD in low or high-grade of IN. 4. We just focus on identifying IN-ECRC from NIN-ECRC, which can’t distinguish between sessile serrated lesions and other lesions. However, about 80% of sporadic CRC develop from IN^18^, removal of which may decrease the incidence of colorectal cancer by 53%. 5. The prospective validation is not a randomized controlled trial (RCT). We’re planning to proceed with subsequent RCT studies in more centers. 6. We just put forward the endoscopic image that can be used to predict molecular subtype, but ECRC-CAD cannot identify surface microstructure yet. Considering the clinical applicability, the model was constructed using images taken by WLE in this study, which were not suitable for micro-structure classification. In subsequent RCT studies, high-resolution WLE and IEE images of early neoplasm will be prospectively collected to construct the classification model for microstructure.

This is an exploratory study on identification and diagnosis of early cancers that could be applied to other organs of digestion system, such as the stomach and esophagus.

## Data Availability

The data would be available upon requests to corresponding authors.

## Acknowledgement

This research was supported by Major Science and Technology Project of Zhejiang Provincial (2020C03030), Natural Science Foundation of Fujian Province (2019-CXB-31), Science and technology program of Guangdong Province (2016A020213002), and National Natural Science Foundation of China (81773956). We would like to thank: (1) Yonghe Zhou, Kunping Hang for coordinating the data in the primary hospitals; (2) Tonghai Dou, Ph.D, Chuncheng Cheng, ZI CHONG KUO providing valuable advice; and Volunteer Joey Yau helps in data collation; (3) The Pucheng county health administration has provided assistance to primary hospitals; (4)The families of all the participants provided spiritual encouragement and material support, allowing researchers to continue their experiments in the most difficult times.

## Contributors

Yiqun Hu is general manager the of the project. Hesong Qiu is the project coordinator and participates in the optimization of research design. Sijun Meng, Wangyue Wang, Yu Zhang, Zhaofang Han, Wenjuan Qin, Yuhuan Zhong, Yi Zhang, Zhenghua Jiang, Yongxiu Zhang, Chao Dong, Yongchao Hu, Lizhong Xie, Jianping Jiang, Huafen Zhu, Wenxia Li, Zhang Wen, Xiaofang Zheng, Yuanlong Sun, Xiaolu Zhou, Limin Ding have contributed in participants’ enrollment, imaging collecting and quality control. Yueping Zheng, Ruizhang Su, Siqi Wu, Yemei Bu, Xiaofang Zheng have classified and integrated data. Hang Xiao and Chen Yang are responsible for algorithm establishing and optimizing. Sijun Meng, Wen Zhang, Lichong Yan, Haineng Xu, and Xugong Li contribute to manuscript writing. Ruizhang Su, Yemei Bu, Wenjuan Qin, Hong Chen, Wensheng Pan, Yiqun Hu were organized and labeled photos. Yiqun Hu, Wensheng Pan, Hong Chen identified the IN-ECRC as expert endoscopist. Shuisheng Wu, Wensheng Pan and Changhua Zhang’s contributed equally in the project.

The corresponding author is Changhua Zhang, Shuisheng Wu, Wensheng Pan, Yiqun Hu. According to the author ranking, Hesong Qiu and the previous author are first co-authors, Xugong Li and the previous author are second co-authors, the others is the third co-authors.

## Ethical approval

This study was approved by the medical ethic committee of each participants (IRB serial number: XMZSYYKY_ER No.2019023).

